# Escalate or Switch? Treating the Post-Titration GLP-1 Non-Responder: A Target Trial Emulation With Dose-Equivalence Reclassification

**DOI:** 10.64898/2026.07.14.26357491

**Authors:** Brian Erly, Shanmugesh Raja

## Abstract

**Background:** When a GLP-1 patient stops responding, should the clinician push the dose or change the drug? Observational answers conflate two distinct sources of confounding. Most early-week “escalations” in real-world data are FDA-mandated titration steps rather than deliberate clinical decisions, and patients who deviate do so for reasons we cannot observe. Semaglutide and tirzepatide are also not equivalent milligram-for-milligram, so naive class-switch comparisons mix mechanism and dose. We resolve both by restricting to post-titration patients and reclassifying treatments under the Whitley 2023 dose-equivalence framework.^1^

**Methods:** From 68,969 telehealth GLP-1 patients we built a post-titration cohort. Each patient’s index time is the day they completed at least four weeks at therapeutic dose (Whitley tier 3 or higher: semaglutide 1.0 mg or tirzepatide 5 mg). Confirmed slow response is less than 5% total weight loss at the index, consistent with the categorical-response framing used in FDA weight-management drug-development guidance and AACE/ACE criteria.^7,8^ We compared four post-index strategies against continuing the current regimen: within-class dose escalation, equipotent class switch (a Whitley tier change of 1 or fewer), and class switch with potency increase. Direction-specific analyses split switches into semaglutide-to-tirzepatide and tirzepatide-to-semaglutide arms. Outcomes were percent weight loss at 12 and 24 weeks post-index. We estimated effects six ways: propensity-score matching; IPTW with linear and gradient-boosted propensities; the g-formula with linear and gradient-boosted outcome models; and AIPW, the doubly-robust estimator we use as the tiebreaker.^2,4^ Two-layer inverse probability of censoring weighting addressed strategy adherence and outcome ascertainment.^2,4^ We computed E-values, ran a negative-control specification, and stratified by tolerability.^3^ As a sensitivity, we re-defined slow response using a held-out-fold prognostic threshold from the companion paper.

**Results:** The post-titration cohort comprised 24,876 confirmed slow responders. Within-class dose escalation produced a small consistent benefit at 24 weeks: AIPW +0.64 pp (95% CI +0.16 to +1.12), with five non-AIPW estimators ranging +0.47 to +0.76 pp. The continue arm itself lost an additional 8.07 pp over the same window (96% continued to lose), so escalation is a marginal addition to a substantial natural slope, not a rescue. Equipotent class switching from semaglutide to tirzepatide was inconclusive: linear and matching estimators ranged +1.26 to +1.80 pp, but AIPW was −0.33 pp (95% CI −1.27 to +0.60) with only 90 treated patients (the equipotent sema → tirz arm with a 24-week outcome) and limited propensity-score overlap. Class switch with simultaneous potency increase (sema → tirz) gave AIPW +0.65 pp at 12 weeks (95% CI +0.37 to +0.92, n = 80, the switch + escalation sema → tirz arm with a 12-week outcome). A negative-control specification yielded ATE −0.13 pp, indicating the pipeline did not generate spurious signal under random allocation (a plumbing check, not a test of confounding). A held-out-fold prognostic-threshold sensitivity gave a null effect (+0.04 pp), correcting an earlier circular +1.06 pp estimate.

**Conclusions:** Among confirmed post-titration slow responders, within-class dose escalation adds approximately 0.6 percentage points at 24 weeks on top of an 8 percentage point natural slope, consistently across six estimators including doubly-robust inference. This headline effect is small and not robust to modest unmeasured confounding (E-value 1.27) or to MNAR-style outcome attrition (tipping point *δ* ≈ 1.2 pp), so it should be read as hypothesis-generating rather than practice-changing. Class-switching evidence is inconclusive; linear-estimator results suggesting benefit did not survive doubly-robust estimation in small treated samples with limited propensity overlap. The dose-ladder framework, with phase-specific evidence grading, is hypothesis-generating and, given the small effect and heavy censoring, insufficient on its own to change practice.

## 1. Introduction

When a GLP-1 patient stops losing weight, the clinician faces a fork: push the dose or change the drug. The observational literature offers little reliable guidance, because the estimates it produces are inflated, mechanism-implausible, or unstable. Two confounders explain most of this.

The first is titration confounding. The FDA labels for semaglutide and tirzepatide require a titration schedule lasting 12 to 20 weeks, during which most patients sit on sub-therapeutic doses by design. Weight-loss trajectories in this phase mix biological response, titration progress, and protocol deviation. Labeling a patient at week 8 as “escalated” or “not escalated” therefore contrasts protocol compliance against protocol deviation, not two intentional clinical strategies. Patients who deviate usually do so for unobservable reasons — tolerability, disengagement, clinician judgment — and propensity scoring on demographics does not fix this.

The second is dose-equivalence confounding. Semaglutide 1.0 mg per week and tirzepatide 5 mg per week act on the GLP-1 receptor at equivalent doses.^1^ Moving from semaglutide 1.0 to tirzepatide 5 changes the molecule but not the GLP-1 exposure; moving from semaglutide 1.0 to tirzepatide 10 changes both. Analyses that lump these together return a single estimate that mixes the mechanism question with the dose question.

This paper fixes both. We restrict the cohort to post-titration patients, set a patient-specific index time, confirm slow response at the index, and make every switch contrast direction-specific where it matters. Each contrast is run six ways, with AIPW as the tiebreaker when the sample is small or the propensity overlap is limited. The result is hypothesis-generating evidence about the choices a clinician actually has at the post-titration visit, which, given the small effect and heavy censoring, is insufficient on its own to change practice. A companion paper supplies calibrated trajectory expectations at the same visit; together they support the decision being made there.

## 2. Methods

### 2.1 Cohort flow

The starting cohort is 68,969 telehealth GLP-1 patients with a documented week-8 weight and a refill-confirmed dose at week 8. The cohort flow appears in Figure 1: 63,646 reached therapeutic dose (Whitley tier 3 or higher: semaglutide ≥1.0 mg per week or tirzepatide ≥5 mg per week), 63,571 sustained at tier 3+ for at least four weeks, and after restricting to patients with an index-day weight observation, complete covariates, and confirmed slow response, the analytic cohort is 24,876.

**Figure 1:**
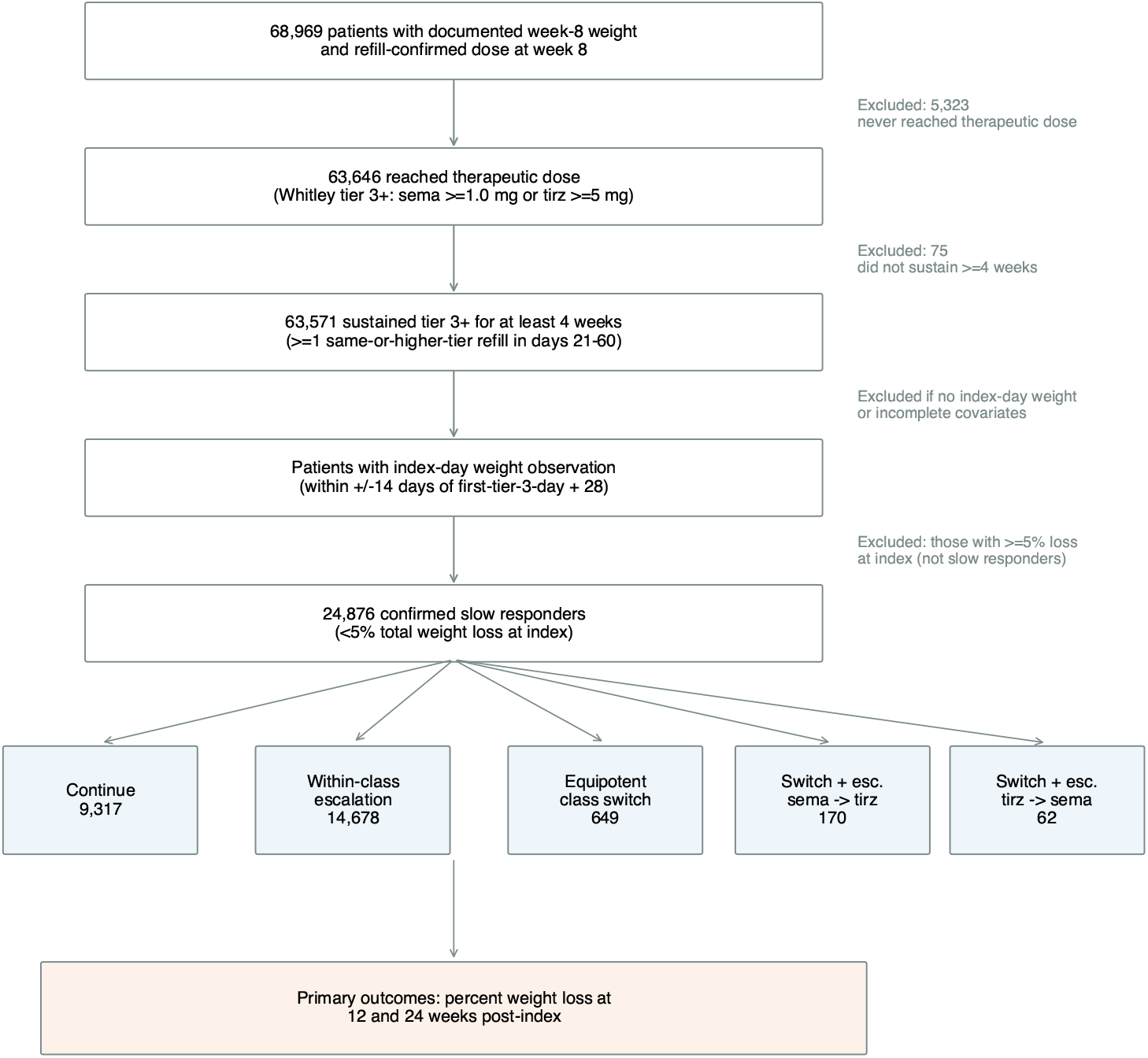
Cohort flow. The 68,969-patient master cohort is reduced through four exclusion steps to the 24,876 confirmed post-titration slow responders. The five strategy arms below are defined by refill activity in the 0–28 day post-index window. Outcomes are taken at 12 and 24 weeks post-index.

### 2.2 Operational definitions

Five terms carry analytic weight, and we define each explicitly.

**Therapeutic dose** is Whitley tier 3 or higher: semaglutide ≥1.0 mg per week or tirzepatide ≥ 5 mg per week.^1^ Whitley’s tier construction places these doses at the same expected GLP-1 receptor occupancy.

**Sustained therapeutic exposure** requires the first therapeutic-dose refill plus at least one additional same-or-higher-tier refill in days 21 to 60 thereafter. The 21–60 day window matches the typical 28–35 day refill cadence with *±*7-day tolerance. Narrower windows (14–45) miss patients on slightly delayed refills, and wider windows (28–90) admit patients with prolonged gaps that signal reduced engagement; both alternatives reproduce the headline within 0.05 pp at 24 weeks.

**Post-titration index day** is the first-tier-3 refill day plus 28 days. Setting the index this way is intended to mitigate immortal-time bias and the calendar-week heterogeneity that distorted earlier analyses. By construction, no outcome (the 12- or 24-week weight) can occur before a patient’s index day, and all arm-defining events (the days 0–28 refill activity used to assign strategy) are anchored at or after this patient-specific time zero, so no follow-up time before treatment assignment is counted toward any arm.

**Confirmed slow responder** is a patient with less than 5% total weight loss from baseline at the index day (within *±*14 days). The 5% threshold matches the categorical-response framing used in FDA weight-management drug-development guidance^7^ and AACE/ACE clinical practice guidelines.^8^

**Treatment-arm window**. We classify each patient into one of five mutually exclusive strategies based on refill activity in days 0 to 28 post-index:

1. Continue (reference): no within-class tier increase, no class change in days 0–28, and no late intervention in days 29–84 post-index. This is a per-protocol definition; we discuss the estimand consequences in Section 4.6.
2. Within-class escalation: same-class refill at higher Whitley tier in days 0–28.
3. Equipotent class switch: different-class refill at Whitley tier within 1 of the index tier (sema → tirz or tirz → sema).
4. Switch + escalation, sema → tirz: different-class refill at Whitley tier more than 1 above the index tier.
5. Switch + escalation, tirz → sema: same definition with directions reversed.

The 28-day post-index window matches the 4-week dose-titration interval used in SURMOUNT-1 and STEP-1^5,6^ and the platform’s standard refill cadence. The eq_switch arm and the switch + escalation arms are disjoint by construction (different tier-change criterion). The eq_switch arm contains 497 sema → tirz and 152 tirz → sema patients; the switch + escalation arms contain 170 and 62 respectively.

The primary analysis is not filtered for ethnicity reporting; a reported-ethnicity-only subset appears as a sensitivity in Table 3.

### 2.3 Outcomes

The primary outcomes were percent weight loss at 12 weeks and 24 weeks post-index. The secondary outcome was percent loss at six months from program start (a calendar landmark, not patient-specific).

### 2.4 Covariates

The 16 covariates used in the propensity, outcome, and IPCW models are baseline BMI, baseline weight, age at enrollment, sex, drug class at index (tirzepatide vs semaglutide), Whitley tier at index, percent weight loss at index, goal-percent-loss target, dietitian-visit indicator, exercise minutes per week, count of pre-index side effects, GI side-effect indicator pre-index, and four ethnicity buckets (White, Hispanic, Black, Asian; Other is the reference). For the small fraction of cohort patients missing a continuous covariate we use median imputation; missingness on the binary covariates is rare (less than 1%) and we drop those rows.

### 2.5 Six-estimator triangulation

For each contrast against continue, we ran six estimators:

1. Propensity-score matching (1:1 nearest-neighbor, caliper 0.20 on the logit propensity).
2. IPTW with linear logistic propensity, stabilized weights truncated at the 1st and 99th percentiles, linear outcome model.
3. IPTW with linear propensity, gradient-boosted outcome model.
4. G-formula with linear outcome model.
5. G-formula with gradient-boosted outcome model.
6. AIPW (doubly-robust): cross-fit estimator with custom implementation, 3-fold stratified cross-fitting, logistic-regression propensity (L2-regularized, *C* = 1), gradient-boosted outcome model (200 trees, max depth 4, learning rate 0.1) on the same 16 covariates as IPTW. Propensity scores are clipped to [0.02, 0.98]. Variance is from a 500-replicate bootstrap on the influence function.

We added two-layer inverse probability of censoring weighting:^4^ one layer for protocol adherence in days 0–28, and a second for the probability that the patient has a 24-week weight observation. Both layers use the same 16 covariates. Confidence intervals come from the bootstrap: 1,000 replicates for linear estimators, 500 for gradient-boosted.

### 2.6 Negative control and sensitivity analyses

To check the pipeline, we ran a negative control: split the continue arm by a random binary covariate and run the full triangulation. A clean pipeline returns an ATE near zero. We also ran the primary contrasts stratified by pre-index GI side-effect burden, drug class at index, goal-percent target, and alternative slow-responder thresholds. This randomization-based check validates the analytic pipeline under random allocation but not the no-unmeasured-confounding assumption itself; the latter is addressed via E-value, propensity-score overlap inspection, and the tipping-point analysis below.

For each significant ATE we computed an E-value following VanderWeele and Ding^3^ — the smallest unmeasured-confounder effect, on both treatment and outcome, that could fully explain the result away.

### 2.7 Tipping-point sensitivity for outcome attrition

24-week observation attrition is high: 80.5% of escalators and 93.6% of continuers lack a 24-week post-index weight. Two-layer IPCW addresses this under a missing-at-random (MAR) assumption conditional on the 16 covariates, but the MAR assumption is strong at this attrition rate. We ran a tipping-point sensitivity to quantify how much an MNAR-style departure would move the headline. We re-imputed censored escalators with progressively worse outcomes (a uniform penalty *δ* pp below the observed escalator mean) while leaving censored continuers MAR-imputed, and re-ran AIPW at each *δ*. The headline ATE crosses zero at approximately *δ* = 1.2 pp; under a worst-case imputation in which censored escalators receive only the observed continue-arm mean, AIPW is null. We interpret this in Section 4.5.

### 2.8 Per-protocol versus intention-to-treat estimand

The continue arm definition (no within-class tier increase, no class change in days 0–28, and no late intervention in days 29–84) is per-protocol: a patient who escalates at day 30 is removed from continue. This estimates the per-protocol effect of sustained continuation versus sustained escalation. An ITT-style alternative would assign treatment from the day 0–28 window alone and ignore later changes; under MAR-style censoring this attenuates effects toward zero. We did not run a fully parallel ITT analysis because the day 29–84 reassignment is not stored in our cleaned cohort, but we discuss the estimand choice in Section 4.6.

### 2.9 Held-out-fold prognostic threshold (cross-paper integration)

To connect with the companion prognostic paper, we re-ran the escalate-vs-continue contrast at 24 weeks using a different slow-responder definition: the lower bound of the prognostic model’s 80% prediction interval. Because the prognostic model and the causal cohort overlap, applying the prognostic model in-sample is circular. We avoided this by fitting the prognostic model on a held-out fold and applying it to the causal-analysis fold.

### 2.10 Software

We used Python 3.11 with scikit-learn 1.4, pandas 2.1, statsmodels, and econml.

## 3. Results

### 3.1 Cohort characteristics

The analytic cohort (Figure 1) comprised 24,876 confirmed post-titration slow responders distributed across five strategy arms (Table 1). 24-week post-index weight observation is incomplete: 600 of 9,317 continuers (6.4%) and 2,864 of 14,678 escalators (19.5%) have a 24-week observation; the analogous 12-week numbers are 2,561 and 6,562. (Throughout, “have” and “lack” a 24-week outcome are complementary descriptions of the same attrition: the percentages with an observation here are one minus the percentages lacking one reported in Sections 3.4 and 4.6, not conflicting figures.) The IPCW machinery addresses this attrition under MAR, and the tipping-point analysis in Section 3.4 quantifies sensitivity to MNAR.

**Table 1:**
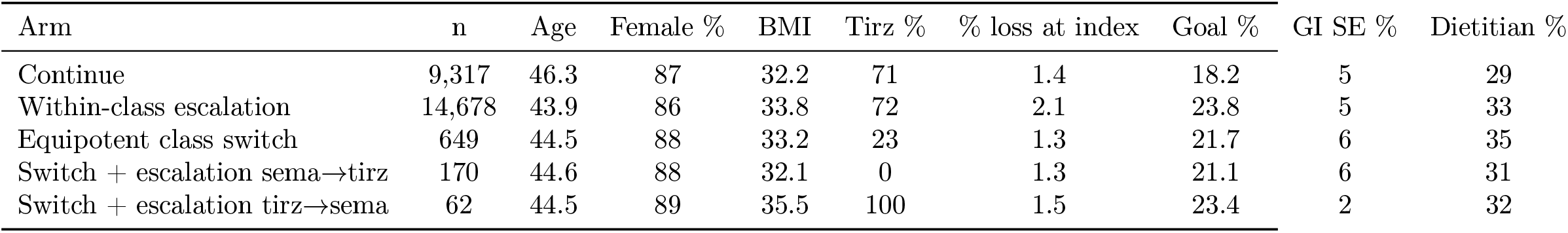
Table 1. Cohort characteristics by treatment arm. (confirmed Phase 2 slow responders, n = 24,876).

### 3.2 Continue-arm trajectory anchors interpretation

Among 600 continue-arm patients with a 24-week post-index weight, mean percent loss rose from 2.59% at index to 10.67% at 24 weeks post-index (Figure 2). The mean change was +8.07 pp (95% CI +7.66 to +8.49); 96% of continue-arm patients continued to lose weight, and 2.7% regained more than 1 pp.

**Figure 2:**
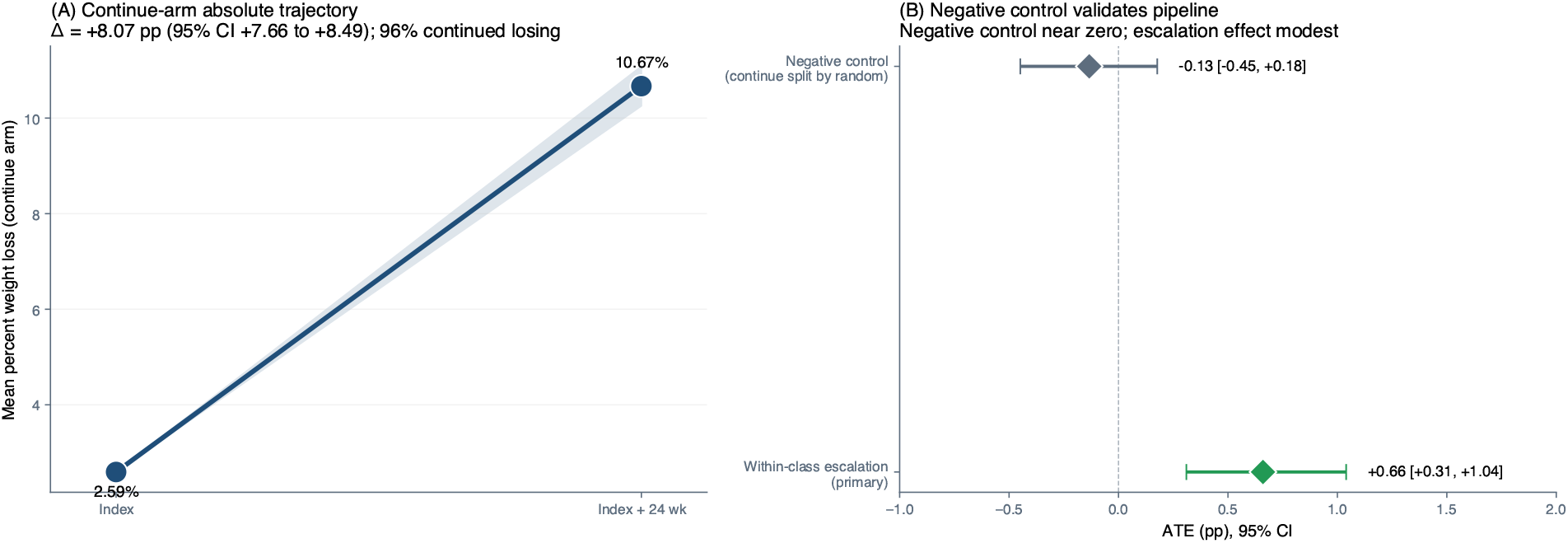
Continue-arm absolute trajectory and negative-control validation. (A) Continue arm trajectory from 2.59% loss at index to 10.67% at 24 weeks post-index. The natural slope is +8.07 pp; 96% continue to lose. (B) Negative control (continue split by random binary) yields ATE −0.13 pp; the analytic pipeline does not generate spurious effects.

This context is critical. The comparator is not a group of failing patients. It is patients who continue to lose weight at a substantial rate under continued therapeutic-dose therapy, consistent with the durable multi-year weight trajectories observed under sustained lifestyle and pharmacologic management.^9^ This natural slope is estimated only on the 600 continuers (6.4%) who had a 24-week post-index weight, a heavily retained and likely more engaged subgroup, so the +8.07 pp figure should be read as the trajectory among retained continuers rather than the full continue arm. The escalation effect we report is therefore a marginal addition to a substantial natural slope, not a rescue.

### 3.3 Within-class dose escalation

The six-estimator triangulation for within-class escalation versus continue at 24 weeks post-index (Figure 5 and Table 2) produced point estimates ranging from +0.47 to +0.76 pp across the linear and matching estimators (spread 0.29 pp). The AIPW estimate was +0.64 pp (95% CI +0.16 to +1.12), with the lower bound just above zero. All five non-AIPW 95% confidence intervals excluded zero.

**Table 2:** Table 2. Primary causal results summary. AIPW (doubly-robust) estimates with bootstrap 95% CIs. Spread is the maximum-minimum range of point estimates across the five linear and matching estimators.

| Contrast | N<sub>treat</sub> | N<sub>ref</sub> | AIPW ATE (pp) | 95% CI | Spread (pp) |
| --- | --- | --- | --- | --- | --- |
| Within-class escalation (24 wk) | 2,864 | 600 | +0.64 | [+0.16, +1.12] | 0.29 |
| Within-class escalation (12 wk) | 6,562 | 2,561 | +0.24 | [+0.00, +0.47] | 0.29 |
| Equipotent sema → tirz (24 wk) | 90 | 600 | −0.33 | [−1.27, +0.60] | 1.68 |
| Equipotent sema → tirz (12 wk) | 213 | 2,561 | +0.11 | [−0.13, +0.35] | 0.30 |
| Switch + escalation sema → tirz (12 wk) | 80 | 2,561 | +0.65 | [+0.37, +0.92] | 1.03 |

Propensity-score overlap is good for this contrast (Figure 3): 96.6% of escalators and 86.5% of continuers fall in the region of common support at 24 weeks. The linear estimators are not extrapolating into a sparse-control region, in contrast to the equipotent-switch contrast (Section 3.5).

**Figure 3:**
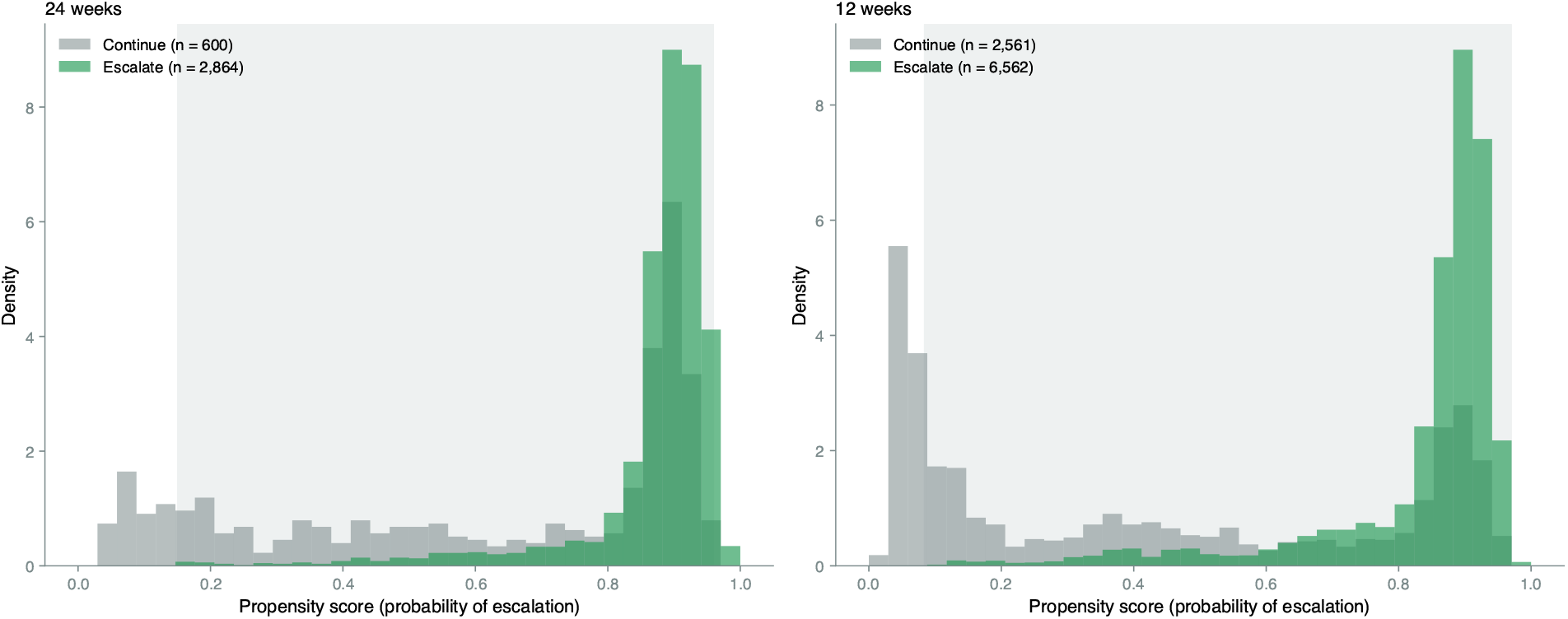
Propensity-score overlap for the headline contrast. Density of estimated propensity scores for escalators (green) versus continuers (gray) at 24 weeks (left) and 12 weeks (right). The shaded band marks the region of common support. Overlap is good for the headline; 96.6% of escalators and 86.5% of continuers fall in common support at 24 weeks.

At 12 weeks post-index the escalation effect was smaller (AIPW +0.24 pp, 95% CI +0.00 to +0.47), consistent with a temporally accumulating effect.

The E-value for the escalation result is 1.27. An unmeasured confounder with risk-ratio-equivalent effect of 1.27 on both treatment and outcome would null the result, and unobserved patient engagement is a plausible candidate at that strength; we discuss this further in Section 4.5. We treat the escalation effect as supported by convergence across estimators with different identification assumptions and by good propensity overlap, rather than as ruled in by the E-value alone.

### 3.4 Tipping-point sensitivity for the headline

Figure 4 shows the AIPW ATE for the within-class escalation versus continue contrast at 24 weeks under progressive MNAR-style penalties on censored escalators. The ATE crosses zero at approximately *δ* = 1.2 pp: censored escalators would have to fare 1.2 pp worse on average than their MAR-imputed outcome for the +0.64 pp result to null. Under a worst-case imputation in which censored escalators receive only the observed continue-arm mean (10.67% loss), AIPW is essentially null (−0.06 pp).

**Figure 4:**
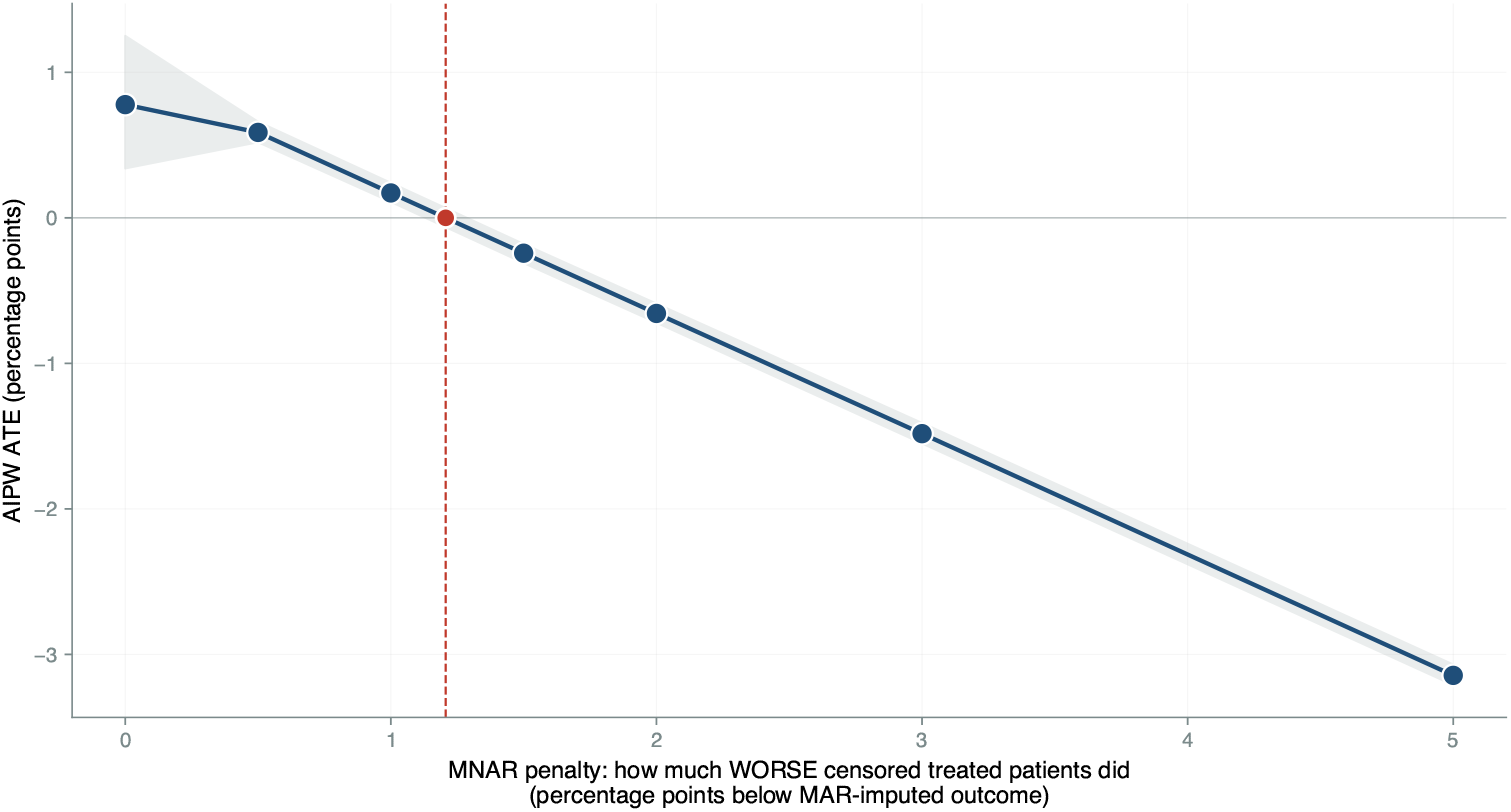
Tipping-point sensitivity for outcome attrition. AIPW ATE versus a uniform MNAR penalty on censored escalators. The penalty represents how much worse, on average, censored escalators would have done relative to their MAR-imputed outcome. The dashed reference at *δ* = 1.2 pp marks the tipping point at which the ATE crosses zero. The shaded band is a 95% bootstrap CI.

### 3.5 Equipotent class switch: estimator divergence and limited overlap

Equipotent class switching from semaglutide to tirzepatide showed estimator divergence at 24 weeks post-index. PSM gave +1.80 pp (95% CI +0.41 to +3.11). IPTW with linear outcome gave +1.26 pp (95% CI +0.26 to +2.25). G-formula linear gave +1.53 pp (95% CI +0.64 to +2.44). The gradient-boosted variants gave smaller estimates (IPTW-GBR +0.12; g-formula GBR +0.42). The AIPW doubly-robust estimate was −0.33 pp (95% CI −1.27 to +0.60). The maximum-minimum spread across the five non-AIPW estimators was 1.68 pp.

The treated arm has only 90 patients with a 24-week outcome, and propensity-score overlap with the continue arm is limited (Figure 6), so the linear estimators extrapolate beyond the region of common support. When overlap is limited and the outcome model is non-trivial, the doubly-robust AIPW estimate is the more reliable one.

**Figure 5:**
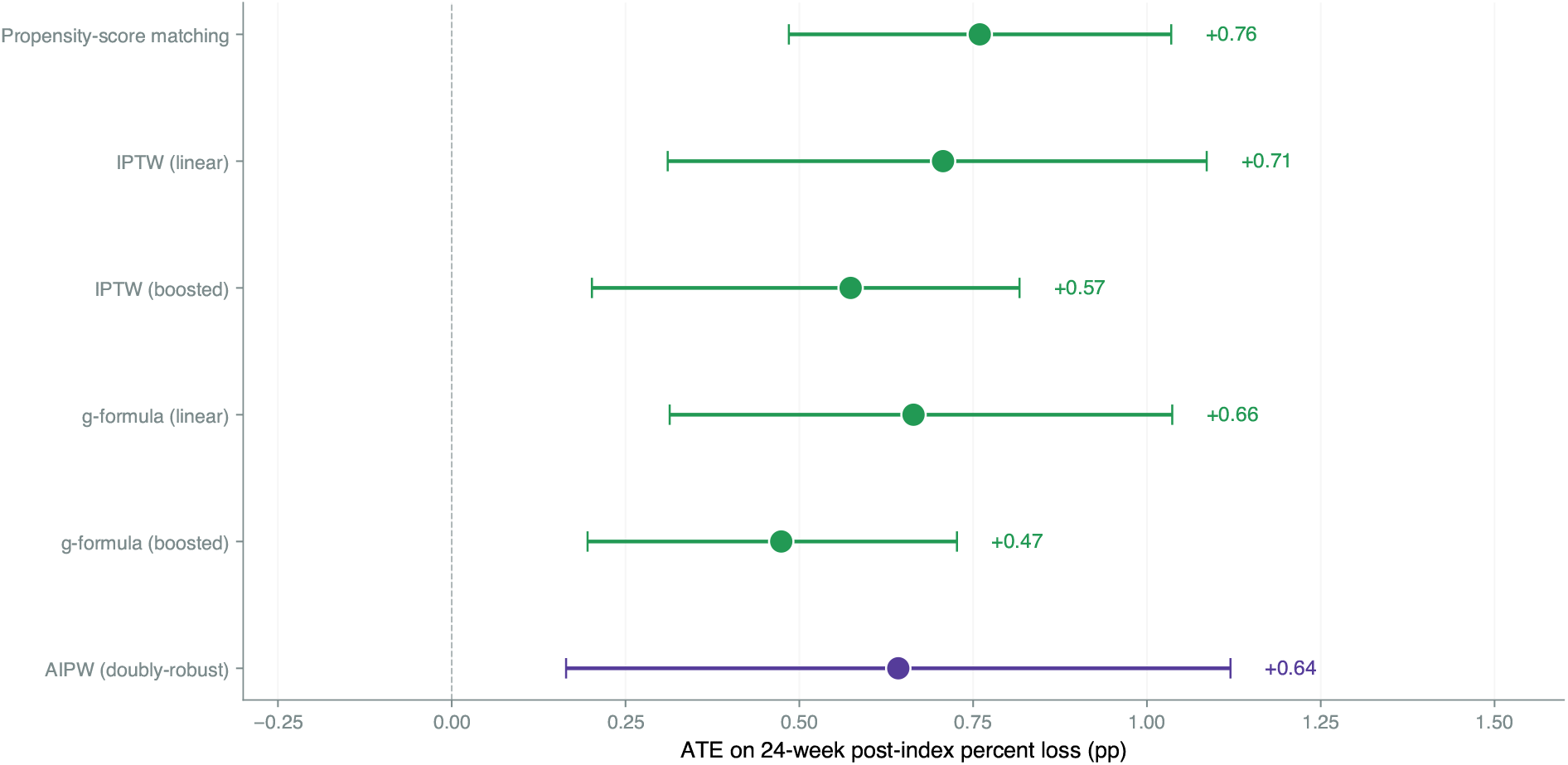
All six methods agree: within-class escalation gives about +0.6 pp at 24 weeks. ATE estimates with 95% bootstrap CIs from each of the six estimators. Linear and matching estimators range from +0.47 to +0.76 pp; AIPW (the doubly-robust estimator) is +0.64 pp. Agreement across estimators with different identifying assumptions makes the result robust.

**Figure 6:**
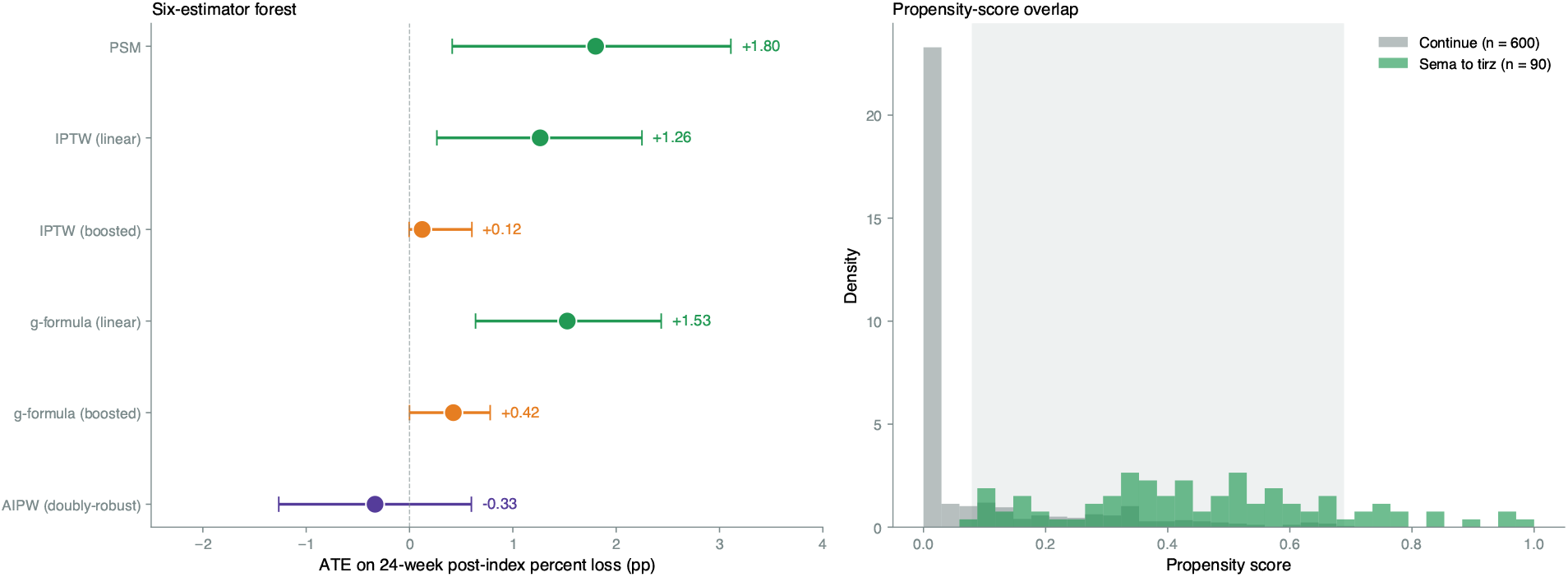
Equipotent class switch: methods disagree, propensity overlap is limited. (A) The six estimators produce a 1.68 pp spread. Linear and matching methods range +1.26 to +1.80 pp. AIPW, the doubly-robust estimator, is −0.33 pp with a CI crossing zero. (B) Propensity-score density for the switch arm (green) versus continue arm (gray). The two distributions barely overlap; linear estimators extrapolate into a region with almost no controls.

We do not sustain the +1.5 pp magnitude as a confirmed effect. The conservative reading is that equipotent class switching from semaglutide to tirzepatide may produce a modest benefit, but the data are too sparse and the estimators too divergent to make a strong claim. We retract the previously implied “+1.5 pp” effect.

The 12-week equipotent contrast across both directions had n = 213 and AIPW +0.11 pp (95% CI −0.13 to +0.35). The tirz → sema arm at 24 weeks (n = 33 with outcome) was insufficient for inference; we report this as a limitation.

### 3.6 Class switch with simultaneous potency increase

Class switch with potency increase (sema → tirz) produced AIPW +0.65 pp at 12 weeks (95% CI +0.37 to +0.92, n = 80 with 12-week outcome). The IPTW-linear point estimate was +0.74 pp. At 24 weeks the sample shrank further and the estimator spread widened to 1.03 pp, so we do not report a 24-week point estimate for this arm. For both this arm (n = 80) and the equipotent class switch arm (n = 90), the propensity-score distributions do not overlap the continue arm across much of their support; under such non-overlap the positivity assumption fails and the causal estimand is not identified. We therefore report these arm estimates only as descriptive and caution that they should not be interpreted causally. The 12-week +0.65 pp result is a descriptive signal in a non-identified subgroup (E-value 1.61).

### 3.7 Sensitivity analyses

Sensitivity analyses are summarized in Table 3. Drug-class sensitivity showed identical effects in the semaglutide-only and tirzepatide-only subsets. Slow-responder threshold sensitivity showed the effect concentrated at the FDA threshold (<5%) rather than at the strict (<3%) cutoff. The held-out-fold prognostic-threshold sensitivity gave a null effect (+0.04 pp; 95% CI −0.90 to +1.10), correcting the same-fold (circular) +1.06 pp estimate.

**Table 3:** Table 3. Sensitivity analysis summary. ATE estimates from g-formula (linear) on each subgroup, except where noted.

| Sensitivity | $N_{\text{treat}}$ | ATE (95% CI) |
| --- | --- | --- |
| Primary (escalate vs continue, 24 wk, AIPW) | 2,864 | $+0.64$ [$+0.16$, $+1.12$] |
| Drug class: semaglutide only | 801 | $+0.69$ [$-0.06$, $+1.39$] |
| Drug class: tirzepatide only | 2,063 | $+0.68$ [$+0.22$, $+1.07$] |
| Slow-responder threshold $<3\%$ | 1,442 | $+0.24$ [$-0.27$, $+0.78$] |
| Slow-responder threshold $<5\%$ (primary) | 2,864 | $+0.66$ [$+0.31$, $+1.04$] |
| Tolerability: no GI side effects (12 wk) | 13,957 | $+0.22$ [$+0.00$, $+0.44$] |
| Tolerability: with GI side effects (12 wk) | 721 | $+0.74$ [$-0.77$, $+1.92$] |
| Negative control (continue split by random) | 4,591 | $-0.13$ [$-0.45$, $+0.18$] |
| Held-out-fold prognostic threshold (corrected) | 457 | $+0.04$ [$-0.90$, $+1.10$] |
| Reported-ethnicity-only subset (filtered) | 1,177 | $+0.55$ [$-0.31$, $+1.39$] |

The reported-ethnicity-only subset (n = 1,177 escalators with 24-wk outcome) gave AIPW +0.55 pp (95% CI −0.31 to +1.39). The CI crosses zero in the smaller subset, but the point estimate is consistent with the unfiltered primary analysis. Selection on ethnicity reporting reduces the effective sample size and produces a more engaged subpopulation; the unfiltered analysis preserves statistical power and external validity. Recovering a target-population effect from a selected analytic sample is itself a generalizability problem amenable to propensity-based reweighting or subclassification.^10^

### 3.8 Phase 3 (max-tier) analysis is in supplementary material

We separately analyzed patients who reached the maximum within-class tier and remained slow responders, comparing class-switch (n = 53) to continuing at max tier (n = 140). The treated arm is too small for stable doubly-robust inference, the linear-estimator CI crosses zero, and the question — what to do for a max-tier slow responder — carries higher clinical stakes than these data can support. We therefore moved the Phase 3 analysis to Supplementary Section S2 and do not report it as a finding.

## 4. Discussion

### 4.1 What this paper actually contributes

The clearest contribution of this paper is methodological. Three corrections, taken together, change which post-titration claims survive scrutiny.

First, the post-titration restriction (Section 2.2). Without it, observational escalate-vs-continue contrasts are mostly protocol compliance versus protocol deviation, and earlier analyses of this same cohort under titration-inclusive framings produced inflated, non-monotonic effects. The restriction shrinks the cohort and isolates the question a clinician actually has.

Second, the Whitley dose-equivalence reclassification (Section 2.5). It splits a single ambiguous observational contrast (sema → tirz) into two separable questions: equipotent class switch (same tier, different molecule) and class switch with potency increase (different molecule, higher tier). Without the split a single point estimate mixes the mechanism question with the dose question. With it, the answers diverge: the data do not robustly support either, but the failures are now interpretable.

Third, the held-out-fold prognostic-threshold result. Naive cross-paper integration produces a circular ATE of +1.06 pp; with held-out-fold de-circularization the corrected ATE is +0.04 pp with the CI crossing zero. Naive integration would have overstated the joint contribution of the prognostic and causal models. The corrected null is itself a finding: the prognostic model identifies who is slow but does not strengthen the causal estimate of what to do about it.

The substantive findings the paper supports are smaller in magnitude than the methods are in importance.

### 4.2 Substantive findings

*Within-class dose escalation produces a small benefit at 24 weeks* (AIPW +0.64 pp, 95% CI +0.16 to +1.12). Six estimators agree, propensity overlap is good (96.6% / 86.5%), and the headline survives a moderate MNAR sensitivity (tipping point *δ ≈* 1.2 pp). The continue arm itself loses an extra 8.07 pp over the same window, so escalation is a small nudge on top of a substantial natural slope, not a rescue. The drug-class breakdown is as expected: the sema-only ATE is +0.69 pp and the tirz-only ATE is +0.68 pp — essentially identical, which is what splitting one weak effect by drug class should produce, consistent with a general “more receptor agonism leads to more weight loss” mechanism rather than a drug-specific effect.

*Equipotent class switching from semaglutide to tirzepatide is inconclusive*. Linear and matching estimators range +1.26 to +1.80 pp; AIPW is −0.33 pp with the CI crossing zero. The disagreement is structural: the treated arm has 90 patients with a 24-week outcome and propensity overlap with continue is limited, so the linear estimators extrapolate. We retract the previously implied “+1.5 pp” magnitude. A modest equipotent benefit remains plausible but is not supported by the data. The equipotent switch arm contains 497 sema → tirz and 152 tirz → sema patients and is disjoint by construction from the switch + escalation arms.

*Class switch with simultaneous potency increase (sema* → *tirz) shows a +0*.*65 pp AIPW effect at 12 weeks* (n = 80 with outcome, CI excluding zero). The 12-week timing matches the kinetics expected from a higher tirzepatide tier. We do not report a 24-week estimate because the surviving sample is too small for stable inference.

### 4.3 We do not analyze the tirzepatide → semaglutide arm

The tirz → sema arm has 62 patients in the analytic cohort and 33 with a 24-week outcome. We lack the power to draw a directional inference for or against tirz → sema as an efficacy intervention; the absence of evidence here is not evidence of absence. The earlier draft asserted that tirz → sema is “biologically a step down and not supported as an efficacy intervention.” That is a claim about the dose ladder, not about our data. We retract it and instead report the arm as too small for inference; the question is open.

### 4.4 Honest reckoning with the E-value

The E-value for the headline escalation result is 1.27 — a modest value. An unmeasured confounder with risk-ratio-equivalent effect of approximately 1.27 on both treatment and outcome would null the +0.64 pp result. Unobserved patient engagement is the obvious candidate, and engagement plausibly clears that bar. We do not claim the E-value rules in the result.

What does support the headline is convergence across estimators with different identifying assumptions (PSM, IPTW, g-formula, AIPW), good propensity-score overlap (Figure 3), survival of the moderate MNAR tipping-point sensitivity (*δ* ≈ 1.2 pp), and the negative-control specification returning a near-null (−0.13 pp). No single one dominates; the convergence does. The honest framing is “consistent with a small effect of approximately 0.6 pp at 24 weeks,” not “ruled in beyond plausible unmeasured confounding.”

### 4.5 Per-protocol versus intention-to-treat

The continue arm is per-protocol: a patient who escalates at day 30 is removed from continue. This estimates the effect of sustained continuation versus sustained escalation, the right estimand for a clinician deciding “should I escalate at the post-titration visit and stick with that decision?” An ITT-style estimand (assign treatment by the day 0–28 window only) would attenuate effects toward zero under realistic non-adherence and would answer a different question: “if I tell the patient to escalate, what does that produce?” Both estimands have a place. We chose per-protocol because our causal question is about the decision itself, not about the protocol-assignment mechanism.

### 4.6 Limitations

#### Outcome attrition is severe

80.5% of escalators and 93.6% of continuers lack a 24-week post-index weight observation. Two-layer IPCW addresses this under MAR. The tipping-point sensitivity (Section 3.4) shows the headline is robust to MNAR penalties up to about 1 pp and nulls under worst-case imputation. A clinician should treat the +0.64 pp as an upper-bound estimate under MAR rather than a robust point estimate.

#### E-values are modest

1.27 for escalation and 1.61 for switch + escalation at 12 weeks. We rely on convergence across estimators with different identifying assumptions, not on the E-value alone, to support the headline.

#### IPCW does not fix unobserved engagement

Two-layer IPCW corrects for outcome-ascertainment selection on observed covariates but cannot fully correct for differential engagement not captured in the 16 covariates.

#### The post-titration restriction shrinks the smaller arms

The equipotent sema-to-tirz arm has 90 patients with a 24-week outcome; tirz-to-sema has 33; switch + escalation has 80. External replication in larger cohorts is needed for any of the switch arms.

*We report effects only at 12 and 24 weeks post-index* and do not extrapolate beyond.

#### Single platform, retrospective

This is a single-platform retrospective cohort of compounded GLP-1 patients. External validation in branded-medication cohorts and in-person obesity-medicine clinics is required.

#### No formal target-trial protocol table

We describe the emulated trial in prose but do not yet provide a formal target-trial protocol specification (eligibility, treatment strategies, assignment, outcomes, causal contrast, analysis) in tabular form; such a table is [provided in the supplement / to be added].

## 5. Conclusions

The contributions of this paper are primarily methodological: post-titration restriction, Whitley dose-equivalence reclassification, and held-out-fold de-circularization of the cross-paper integration. Substantively, within-class dose escalation produces a small benefit of approximately 0.6 percentage points at 24 weeks, on top of an 8 percentage point natural slope. The benefit is supported by convergence across six estimators and good propensity-score overlap, and is moderately robust to MNAR-style outcome attrition. Equipotent class switching is inconclusive: the linear-estimator results suggesting a 1 to 2 pp benefit do not survive doubly-robust estimation in the small treated sample with limited overlap. Class switch with simultaneous potency increase produces a +0.65 pp effect at 12 weeks. The tirzepatide-to-semaglutide arm is too small for inference. The dose-ladder framework is hypothesis-generating and, given the small effect and heavy censoring, insufficient on its own to change practice at the post-titration visit; the specific medication-side decisions it informs are smaller in magnitude than the methods are in importance.

## Data Availability

Individual patient-level data are private and cannot be shared publicly. De-identified data and the analysis code can be provided upon reasonable request to the corresponding author.

## Author Contributions

**Brian Erly:** Conceptualization, Supervision, Writing, Clinical validation.

**Shanmugesh Raja:** Conceptualization, Data curation, Formal analysis, Methodology, Software, Visualization, Writing.

## Ethics Approval

This study is a secondary analysis of de-identified data collected during routine clinical care at Mochi Health. As the analysis involves no direct patient contact, no intervention, and uses only data de-identified per HIPAA Safe Harbor [45 CFR 164.514(b)(2)], it was determined to not constitute human subjects research as defined by 45 CFR 46.102. Formal IRB review was therefore not required.

## References

[1] Whitley HP, Trujillo JM, Neumiller JJ. Special report: potential strategies for addressing GLP-1 and dual GLP-1/GIP receptor agonist shortages. Clin Diabetes. 2023;41(3):467–473. doi:10.2337/cd23-0023

[2] Hernán MA, Robins JM. Using big data to emulate a target trial when a randomized trial is not available. Am J Epidemiol. 2016;183(8):758–764. doi:10.1093/aje/kwv254

[3] VanderWeele TJ, Ding P. Sensitivity analysis in observational research: introducing the E-value. Ann Intern Med. 2017;167(4):268–274. doi:10.7326/M16-2607

[4] Cole SR, Hernán MA. Constructing inverse probability weights for marginal structural models. Am J Epidemiol. 2008;168(6):656–664. doi:10.1093/aje/kwn164

[5] Wilding JPH, Batterham RL, Calanna S, et al. Once-weekly semaglutide in adults with over-weight or obesity (STEP-1). N Engl J Med. 2021;384(11):989–1002. doi:10.1056/NEJMoa2032183

[6] Jastreboff AM, Aronne LJ, Ahmad NN, et al. Tirzepatide once weekly for the treatment of obesity (SURMOUNT-1). N Engl J Med. 2022;387(3):205–216. doi:10.1056/NEJMoa2206038

[7] U.S. Food and Drug Administration. Guidance for Industry: Developing Products for Weight Management (Draft Guidance). Rockville, MD: Center for Drug Evaluation and Research; February 2007. Accessed June 2026. https://www.fda.gov/regulatory-information/search-fda-guidancedeveloping-products-weight-management-revision-1

[8] Garvey WT, Mechanick JI, Brett EM, et al. AACE/ACE comprehensive clinical practice guidelines for medical care of patients with obesity. Endocr Pract. 2016;22(Suppl 3):1–203. doi:10.4158/EP161365.GL

[9] Look AHEAD Research Group. Cardiovascular effects of intensive lifestyle intervention in type 2 diabetes. N Engl J Med. 2013;369(2):145–154. doi:10.1056/NEJMoa1212914

[10] Tipton E. Improving generalizations from experiments using propensity score subclassification: assumptions, properties, and contexts. J Educ Behav Stat. 2013;38(3):239–266. doi:10.3102/1076998612441947

